# Aeromedical Retrieval of Critically Ill Pulmonary Embolism Patients: A Protocol for a Retrospective Cohort Study of 10 Years in New South Wales

**DOI:** 10.1101/2024.11.02.24316651

**Authors:** Ruan Vlok, Yousif Rassam, Christopher Partyka

## Abstract

**Background:** Pulmonary embolism is a common, time critical condition care requiring multidisciplinary care. Interhospital transport is a high-risk period in the patients’ care, but offers an opportunity for expedited risk stratification, resuscitation and transport to definitive care. Patients who require interhospital transfer to specialist centres for pulmonary embolism management have worse outcomes. Despite this, literature surrounding the medical retrieval practice and experience of pulmonary embolism is limited.

**Methods:** A retrospective cohort study over a 10-year period from January 2014-September 2024 from the database of a high-volume aeromedical retrieval service in New South Wales, Australia will be performed. The study aims to describe the experience in this service in the care of critically ill pulmonary embolism patients requiring interhospital transport.

## Background

Pulmonary embolism (PE) is a common and life-threatening condition. PE has had 30-day mortality of 17.6%-26.5% reported irrespective of severity of initial illness [1]. In Australia, PE and deep vein thrombosis (DVT) has an annual incidence of 0.83 per 1000 population, with an estimated cost of $1.7 billion to the Australian community in 2008 [2].

Treatment modalities for PE have expanded rapidly in recent years, ranging from anticoagulation to catheter-based therapies, surgical embolectomy, extracorporeal membrane oxygenation (ECMO) and systemic thrombolysis. Risk stratification is essential in guiding the need for increasingly invasive interventions. Guidelines differ with regards to optimal risk stratification, particularly in the submassive or intermediate cohort, where treatment may differ significantly [3,4]. This increased complexity in what is frequently a time critical condition has led to the introduction of Pulmonary Embolism Response Teams (PERT) in the United States [5]. These teams serve to allow key senior decision makers, including respiratory physicians, emergency physicians, intensivists, haematologists, interventional radiologists, cardiac surgeons and cardiologists to guide care in a rapid and coordinated fashion. Observational data from established services imply a mortality benefit and reduced hospital length of stay associated with well-coordinated PERT teams. These systems have recently begun to be implemented in Australia, with individualised approaches for a large metropolitan Sydney hospital [6] and a regional Queensland hospital [7].

The PERT Consortium has identified the coordination and care of patients with PE requiring interhospital transfer as a high-risk period and an area in need of research [8]. Patients requiring interhospital transfer for the management of their PE are recognised to have a higher mortality than patients that present directly to specialist centres [9]. To date, to the best of our knowledge, no literature exists regarding the clinical experience of the management of critically ill PE patients during interhospital transfer by medical retrieval services. In New South Wales (NSW), aeromedical retrieval teams (NSWA-AO) transfer patients from a wide range of care facilities across the state to definitive care, including rural, regional and metropolitan interhospital transfers. These referring centres have a wide range of clinical capacities, ranging from rural nurse run clinics to metropolitan intensive care units. This service is frequently the first point of senior critical care contact a patient may have during their healthcare presentation, and therefore represents an opportunity to initiate earlier risk stratification, resuscitation and coordination of PE care, whilst aiming to provide safe transport to definitive care at an appropriate receiving centre. Transport distances by the service varies greatly.

Given the lack of current available literature, and variations in published guidelines, this study will aim to describe the clinical experience of three NSW Aeromedical Retrieval Service bases over the preceding 10 years in caring for critically ill PE patients during their interhospital retrieval.

## Methodology

This manuscript outlines the protocol for a multi-base retrospective cohort study of NSW Ambulance aeromedical database, AirMaestro (Avinet, Australia), from January 2014 to September 2024. This study will be performed in accordance with the STROBE guidelines [10].

### Primary Objective

1. A descriptive summary of all patients retrieved by NSWA-AO for the primary reason of pulmonary embolism.

### Secondary Objectives

2 Describe the risk stratification of patients with PE requiring intrahospital transport according to the ESC classification of PE severity.
3 Describe the pretransfer optimisation provided in patients with PE including anticoagulation, thrombolysis, respiratory support, cardiovascular support and general supportive critical care prior to departure.
4 Describe the characteristics of referring and receiving hospitals involved in the interhospital transfer of patients with PE.
5 Described the post-retrieval management of these patients, including the need for systemic thrombolysis, ECMO support, interventional radiology-based therapies (e.g., catheter directed thrombolysis, catheter embolectomy, IVC filter) or surgical embolectomy.
6 Describe the complications encountered by the medical retrieval team transporting pulmonary embolism patients, including cardiac arrest, bleeding complications, clinically significant hypotension and clinically significant hypoxia.

### Inclusion Criteria

1. Pulmonary embolism for interhospital transfer requiring aeromedical retrieval between January 2014-September 2024.
2. Retrieval facilitated by Sydney, Orange and Wollongong aeromedical bases.
3. Pulmonary embolism representing the primary indication for interhospital transfer, or a diagnosis of acute pulmonary embolism complicating the treatment in an alternative diagnosis.

### Exclusion Criteria

1. History of pulmonary embolism not currently contributing to the decision to transfer the patient
2. Septic pulmonary embolism because of likely infective endocarditis, being transferred for the primary indication of management of infective endocarditis.
3. Low risk pulmonary embolism complicating an alternative diagnosis with no influence on the care required by the patient.
4. Retrieval facilitated by bases other than Sydney, Wollongong or Orange.
5. Prehospital cases of pulmonary embolism responded to by medical retrieval team.

### Ethics

This study was granted ethics approval by the South Western Sydney Local Health District Human Research Ethics Committee (SWSLHD HREC). Ethics approval was granted on 1/11/2024 for as Low and Negligible Risk research with a waiver of consent (2024/ETH02380). A waiver of participant consent was sought as this study is retrospective and non-invasive in that it uses demographic, physiological and patient treatment records which is measured routinely as part of clinical care and analyses the predictive power of these data in relation to later outcomes. This data will not be changed or used in any other way than for the purpose of this study. No other data on the recruited patients will be collected in addition to that used in routine patient assessment and management and routine 24 hours follow up. Once clinical data collected, the patients will be de-identified, and data stored with patients allocated a unique study number.

There is no risk to the rights, privacy or professional reputation of carers, health professionals and/or institutions as the study concerns the care of a common clinical presentation with recognised complexity and common treatment pathways and has no intent to identify individual clinicians or carers, nor to use the data as commentary on the institutions concerned. There is no specific participation required by participants. With patient data being completely de-identified during the analysis there are no specific safety considerations to participants.

### Data Collection

The AirMaestro database will be searched from January 2014 to September 2024. This database will be searched for all cases that were classified by the diagnostic category ‘Pulmonary Embolism’. A secondary search of the database will be performed for any cases including the phrases “pulmonary embolus”, “PE”, “thrombolysis”, and “submassive”. Results will be screened for inclusion and exclusion criteria prior to data extraction.

Anonymised data will be extracted from Air Maestro directly into RedCAP [11,12], including utilising contemporaneous notes, any scanned documents such as observation strips or case sheets, and 24hr follow up on Air Maestro. Table 1 represents the Data Extraction Form, and the ability to add free text will be available for each variable. Where free text is added, two authors will independently review this free text to ensure the extracted variable is accurate and represents the intended data.

**Table 1.**
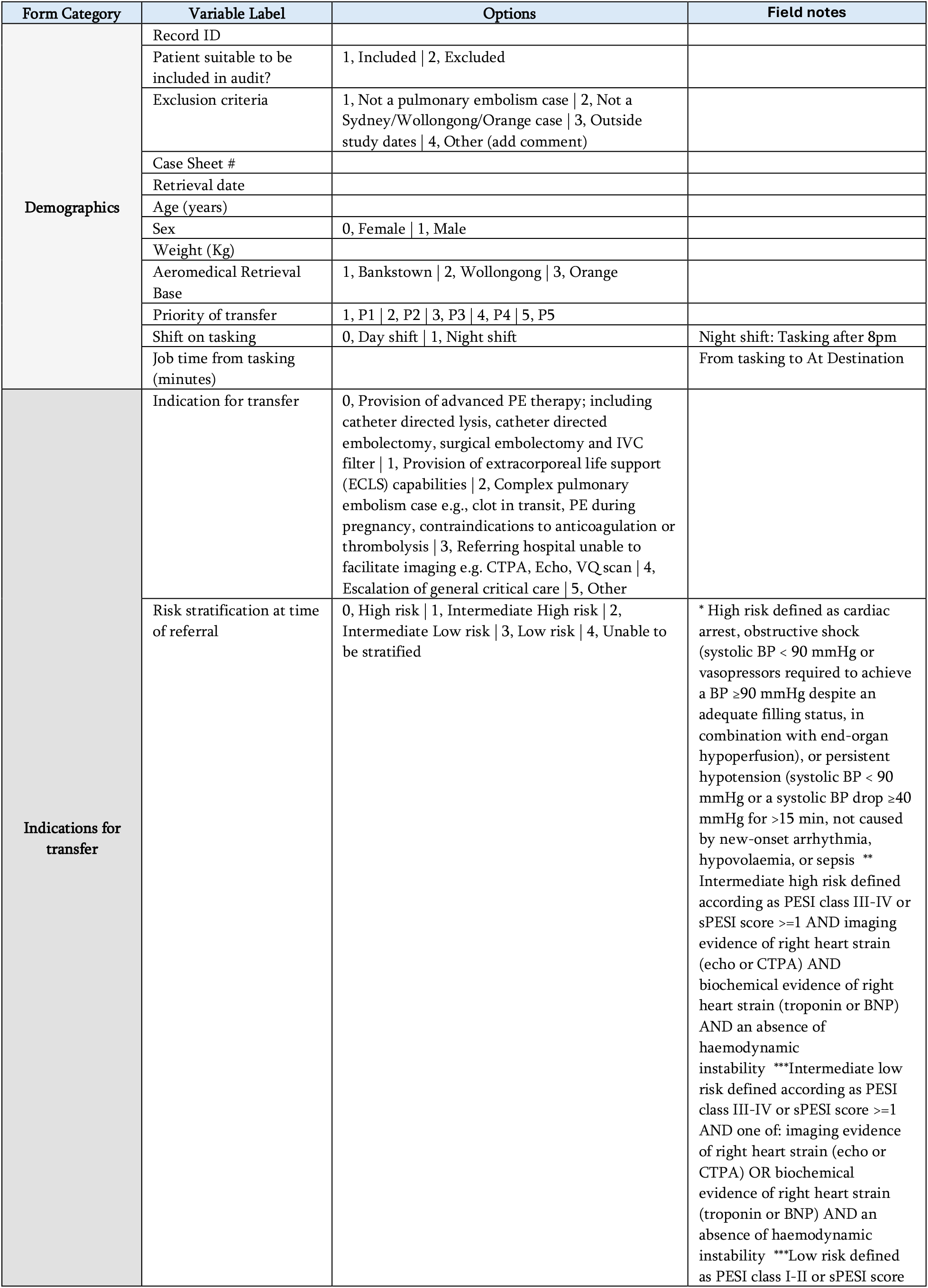
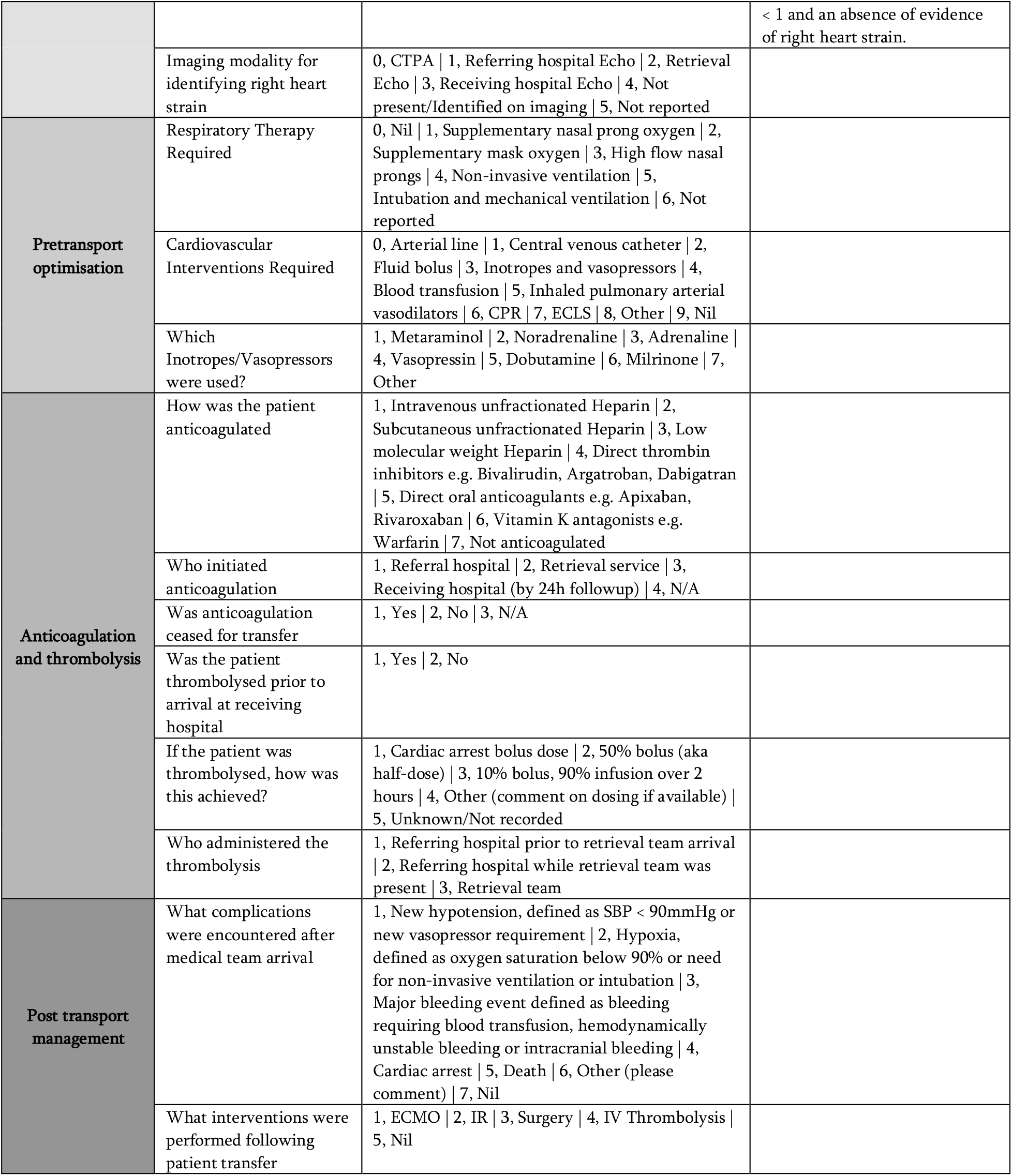
Data Extraction Form.

| Form Category | Variable Label | Options | Field notes |
| --- | --- | --- | --- |
| Demographics | Record ID |  |  |
|  | Patient suitable to be included in audit? | 1, Included \| 2, Excluded |  |
|  | Exclusion criteria | 1, Not a pulmonary embolism case \| 2, Not a Sydney/Wollongong/Orange case \| 3, Outside study dates \| 4, Other (add comment) |  |
|  | Case Sheet # |  |  |
|  | Retrieval date |  |  |
|  | Age (years) |  |  |
|  | Sex | 0, Female \| 1, Male |  |
|  | Weight (Kg) |  |  |
|  | Aeromedical Retrieval Base | 1, Bankstown \| 2, Wollongong \| 3, Orange |  |
|  | Priority of transfer | 1, P1 \| 2, P2 \| 3, P3 \| 4, P4 \| 5, P5 |  |
|  | Shift on tasking | 0, Day shift \| 1, Night shift | Night shift: Tasking after 8pm |
|  | Job time from tasking (minutes) |  | From tasking to At Destination |
| Indications for transfer | Indication for transfer | 0, Provision of advanced PE therapy; including catheter directed lysis, catheter directed embolectomy, surgical embolectomy and IVC filter \| 1, Provision of extracorporeal life support (ECLS) capabilities \| 2, Complex pulmonary embolism case e.g., clot in transit, PE during pregnancy, contraindications to anticoagulation or thrombolysis \| 3, Referring hospital unable to facilitate imaging e.g. CTPA, Echo, VQ scan \| 4, Escalation of general critical care \| 5, Other |  |
|  | Risk stratification at time of referral | 0, High risk \| 1, Intermediate High risk \| 2, Intermediate Low risk \| 3, Low risk \| 4, Unable to be stratified | * High risk defined as cardiac arrest, obstructive shock (systolic BP < 90 mmHg or vasopressors required to achieve a BP ≥90 mmHg despite an adequate filling status, in combination with end-organ hypoperfusion), or persistent hypotension (systolic BP < 90 mmHg or a systolic BP drop ≥40 mmHg for >15 min, not caused by new-onset arrhythmia, hypovolaemia, or sepsis **<br>Intermediate high risk defined according as PESI class III-IV or sPESI score ≥1 AND imaging evidence of right heart strain (echo or CTPA) AND biochemical evidence of right heart strain (troponin or BNP) AND an absence of haemodynamic instability ***Intermediate low risk defined according as PESI class III-IV or sPESI score ≥1 AND one of: imaging evidence of right heart strain (echo or CTPA) OR biochemical evidence of right heart strain (troponin or BNP) AND an absence of haemodynamic instability ***Low risk defined as PESI class I-II or sPESI score |
|  |  |  | < 1 and an absence of evidence of right heart strain. |
|  | Imaging modality for identifying right heart strain | 0, CTPA \| 1, Referring hospital Echo \| 2, Retrieval Echo \| 3, Receiving hospital Echo \| 4, Not present/Identified on imaging \| 5, Not reported |  |
| Pretransport optimisation | Respiratory Therapy Required | 0, Nil \| 1, Supplementary nasal prong oxygen \| 2, Supplementary mask oxygen \| 3, High flow nasal prongs \| 4, Non-invasive ventilation \| 5, Intubation and mechanical ventilation \| 6, Not reported |  |
|  | Cardiovascular Interventions Required | 0, Arterial line \| 1, Central venous catheter \| 2, Fluid bolus \| 3, Inotropes and vasopressors \| 4, Blood transfusion \| 5, Inhaled pulmonary arterial vasodilators \| 6, CPR \| 7, ECLS \| 8, Other \| 9, Nil |  |
|  | Which Inotropes/Vasopressors were used? | 1, Metaraminol \| 2, Noradrenaline \| 3, Adrenaline \| 4, Vasopressin \| 5, Dobutamine \| 6, Milrinone \| 7, Other |  |
| Anticoagulation and thrombolysis | How was the patient anticoagulated | 1, Intravenous unfractionated Heparin \| 2, Subcutaneous unfractionated Heparin \| 3, Low molecular weight Heparin \| 4, Direct thrombin inhibitors e.g. Bivalirudin, Argatroban, Dabigatran \| 5, Direct oral anticoagulants e.g. Apixaban, Rivaroxaban \| 6, Vitamin K antagonists e.g. Warfarin \| 7, Not anticoagulated |  |
|  | Who initiated anticoagulation | 1, Referral hospital \| 2, Retrieval service \| 3, Receiving hospital (by 24h followup) \| 4, N/A |  |
|  | Was anticoagulation ceased for transfer | 1, Yes \| 2, No \| 3, N/A |  |
|  | Was the patient thrombolysed prior to arrival at receiving hospital | 1, Yes \| 2, No |  |
|  | If the patient was thrombolysed, how was this achieved? | 1, Cardiac arrest bolus dose \| 2, 50% bolus (aka half-dose) \| 3, 10% bolus, 90% infusion over 2 hours \| 4, Other (comment on dosing if available) \| 5, Unknown/Not recorded |  |
|  | Who administered the thrombolysis | 1, Referring hospital prior to retrieval team arrival \| 2, Referring hospital while retrieval team was present \| 3, Retrieval team |  |
| Post transport management | What complications were encountered after medical team arrival | 1, New hypotension, defined as SBP < 90mmHg or new vasopressor requirement \| 2, Hypoxia, defined as oxygen saturation below 90% or need for non-invasive ventilation or intubation \| 3, Major bleeding event defined as bleeding requiring blood transfusion, hemodynamically unstable bleeding or intracranial bleeding \| 4, Cardiac arrest \| 5, Death \| 6, Other (please comment) \| 7, Nil |  |
|  | What interventions were performed following patient transfer | 1, ECMO \| 2, IR \| 3, Surgery \| 4, IV Thrombolysis \| 5, Nil |  |

All data will be entered and stored using REDCap software. REDCap is a data collection and storage software hosted on the NSW Ambulance internal server and not accessible to persons outside of the organisation. REDCap meets the United States Health Insurance Portability and Accountability Act of 1996 security requirements. REDCap also has a file repository. Any paper records generated by this study will be scanned and uploaded to the REDCap software and hardcopies will be kept in a secure filing cabinet in the Research Governance office. All electronic data and hardcopy files will be kept for a minimum of seven years and then destroyed securely. At the time of writing this protocol there is no foreseen secondary use of these data following study completion.

## Data Availability

This is a protocol and as such no data is available.

## Data Processing and Analysis

Data analysis will be limited to simple descriptive and frequentist statistics and performed using STATA 18 [13]. Baseline characteristics and demographics were described using median and interquartile range (IQR) for non-normally distributed continuous and ordinal variables. Counts and proportions were presented for dichotomous variables.

## References

1. Acute pulmonary embolism: clinical outcomes in the International Cooperative Pulmonary Embolism Registry (ICOPER) Goldhaber, Samuel Z et al. The Lancet, Volume 353, Issue 9162, 138–1389; Puurunen MK, Gona PN, Larson MG, Murabito JM, Magnani JW, O’Donnell CJ. Epidemiology of venous thromboembolism in the Framingham Heart Study. Thromb Res 2016; 145: 27–33.]

2. Ho WK, Hankey GJ, Eikelboom JW. The incidence of venous thromboembolism: a prospective, community-based study in Perth, Western Australia. Med J Aust. 2008;189(3):144–147

3. Jaff MR, McMurtry MS, Archer SL, Cushman M, Goldenberg N, Goldhaber SZ, Jenkins JS, Kline JA, Michaels AD, Thistlethwaite P, Vedantham S, White RJ, Zierler BK; on behalf of the American Heart Association Council on Cardiopulmonary, Critical Care, Perioperative and Protocol: Interhospital Retrieval of PE v1.01 (15/10/2024) 9 Resuscitation; American Heart Association Council on Peripheral Vascular Disease; American Heart Association Council on Arteriosclerosis, Thrombosis and Vascular Biology. Management of massive and submassive pulmonary embolism, iliofemoral deep vein thrombosis, and chronic thromboembolic pulmonary hypertension: a scientific statement from the American Heart Association [published corrections appear in Circulation. 2012;125:e496 and Circulation. 2012;126:e495]. Circulation. 2011;123:1788–1830. doi:10.1161/CIR.0b013e318214914f

4. Konstantinides SV. 2014 ESC guidelines on the diagnosis and management of acute pulmonary embolism. Eur Heart J. 2014;35:3145–3146. doi: 10.1093/eurheartj/ehu393

5. Barnes GD, Kabrhel C, Courtney DM, Naydenov S, Wood T, Rosovsky R, Rosenfield K, Giri J; National PERT Consortium Research Committee. Diversity in the pulmonary embolism response team model: an organizational survey of the National PERT Consortium members. Chest. 2016;150:1414–1417. doi: 10.1016/j.chest.2016.09.034

6. Roy B, Cho JG, Baker L, Thomas L, Curnow J, Harvey JJ, Geenty P, Banerjee A, Lai K, Vicaretti M, Erksine O, Li J, Alasady R, Wong V, Tai JE, Thirunavukarasu C, Haque I, Chien J; Westmead Hospital Pulmonary Embolism Response Team. Pulmonary embolism response teams. A description of the first 36-month Australian experience. Intern Med J. 2024 Aug;54(8):1283–1291.

7. The Use of a Pulmonary Embolism Response Team (PERT) and Catheter-Directed Treatment in a Far North Queensland Regional Centre Pleash, J. et al. Heart, Lung and Circulation, Volume 32, S453 – S454.

8. Rali, P., Sacher, D., Rivera-Lebron, B., Rosovsky, R., Elwing, J. M., Berkowitz, J., … & Ross, C. B. (2021). Interhospital transfer of patients with acute pulmonary embolism: challenges and opportunities. Chest, 160(5), 1844–1852.

9. Carroll, B. J., Beyer, S. E., Shanafelt, C., Kabrhel, C., Rali, P., Rivera-Lebron, B., … & Secemsky, E. A. (2022). Interhospital transfer for the management of acute pulmonary embolism. The American Journal of Medicine, 135(4), 531–535

10. Von Elm, E., Altman, D. G., Egger, M., Pocock, S. J., Gøtzsche, P. C., & Vandenbroucke, J. P. (2007). The Strengthening the Reporting of Observational Studies in Epidemiology (STROBE) statement: guidelines for reporting observational studies. The lancet, 370(9596), 1453–1457.

11. Harris, P. A., Taylor, R., Thielke, R., Payne, J., Gonzalez, N., & Conde, J. G. (2009). Research electronic data capture (REDCap)—a metadata-driven methodology and workflow process for providing translational research informatics support. Journal of biomedical informatics, 42(2), 377–381.

12. Harris, P. A., Taylor, R., Minor, B. L., Elliott, V., Fernandez, M., O’Neal, L., … & REDCap Consortium. (2019). The REDCap consortium: building an international community of software platform partners. Journal of biomedical informatics, 95, 103208.

13. StataCorp. 2023. Stata Statistical Software: Release 18. College Station, TX: StataCorp LLC.

